# The Remote Assessment of Parkinsonism Supporting Ongoing Development of Interventions in Gaucher Disease – Study Protocol

**DOI:** 10.1101/2021.07.21.21260533

**Authors:** Abigail Louise Higgins, Marco Toffoli, Stephen Mullin, Chiao-Yin Lee, Sofia Koletsi, Micol Avenali, Fabio Blandini, Anthony H V Schapira

## Abstract

Mutations in GBA which are causative of Gaucher disease in their biallelic form, are the most common genetic risk factor for Parkinson disease. The diagnosis of Parkinson disease relies upon clinically defined motor features which appear after irreversible neurodegeneration. Prodromal symptoms of Parkinson disease may provide a means to predict latent pathology, years before the onset of motor features. Previous work has reported prodromal features of Parkinson disease in GBA mutation carriers, however this has been insufficiently sensitive to identify those that will develop Parkinson disease. The Remote Assessment of Parkinsonism Supporting Ongoing Development of Interventions in Gaucher Disease (RAPSODI) study assesses a large cohort of GBA mutation carriers, to aid development of procedures for earlier diagnosis of Parkinson disease.

**Summary Points:**

- The therapeutic focus for Parkinson disease is currently symptomatic, rather than disease-modifying or neuroprotective.
- Non-motor prodromal features of Parkinson disease may precede the motor symptoms required for clinical diagnosis by several years.
- *GBA* mutations, which are associated with Gaucher disease in their biallelic form, have emerged as the most common genetic risk factor for Parkinson Disease.
- *GBA* associated PD displays a slightly exaggerated phenotype, including earlier age of onset, enhanced cognitive decline, more severe affective disturbances, and a greater likelihood of manifesting REM Sleep Behaviour Disorder, hyposmia and autonomic dysfunction.
- The RAPSODI GD study is an annual remote assessment which aims to define the clinical prodrome of PD in a large cohort of *GBA* mutation carriers, to accurately predict clinical diagnosis.
- The secondary objectives of RAPSODI GD are to risk stratify homozygote and heterozygote *GBA* mutation carriers for PD risk, understand the variable penetrance of the *GBA* associated PD phenotype, and create a platform for the future evaluation of biomarkers of disease.
- The putative implication of the RAPSODI GD study is to contribute towards earlier diagnosis of *GBA* associated PD, to provide a timeframe for delivering neuroprotective interventions.
- Defining prodromal PD in *GBA* carriers may have wider implications for sporadic PD.

## Background

### Challenges facing early diagnosis and intervention in Parkinson disease

Parkinson Disease (PD) is the second most common neurodegenerative disease, with increasing global prevalence which doubled between 1990 and 2016 [1,2]. The clinicopathology of PD is characterised by motor symptoms including bradykinesia, tremor and rigidity, attributed to the progressive degeneration of dopaminergic neurons in the substantia nigra pars compacta, [3,4]. An important neuropathological hallmark of PD is the presence of filamentous intracellular inclusions of α-synuclein, known as Lewy Bodies [5].

Current treatments of PD focus primarily on symptom management, rather than disease-modification [6]. With an aging population, the putative continuation of the upward trend in cases of PD calls for the development of preventative therapies, of which there currently are none [7,8]. One of the main challenges for developing neuroprotective therapies for PD is that clinical diagnosis relies upon the presentation of defined motor deficits, which manifest after the onset of neurodegeneration [9]. As such, there is increasing interest in characterising the prodromal features of PD, which may precede motor symptoms by several years [10]. Several latent features of PD’s clinical prodrome have been validated thus far, including REM sleep behaviour disorder (RBD), hyposmia, depression and anxiety, global cognitive impairment, constipation and autonomic dysfunction [11]. In recent years, studies such as PREDICT-PD, have optimised the validation of prodromal features of PD through remote assessment methods, enhancing recruitment and power, and allowing for repeated measures without inter-observer variation [12].

### Gaucher disease and *GBA*

The *GBA* gene located on chromosome 1q21, encodes for the lysosomal hydrolase glucocerebrosidase (GCase), and has 12 exons (exon 1 is noncoding) [13,14]. Gaucher disease (GD) is a rare autosomal recessive disorder caused by biallelic homozygous or compound heterozygous *GBA* mutations, of which there have been over 400 pathogenic mutations identified [15,16,17]. In GD, a loss of function of *GBA* results in deficient GCase activity, and the subsequent accumulation of its substrate, a sphingolipid called glucosylceramide primarily in macrophages [18]. Literature concerning the epidemiology of GD is highly variable, with birth incidence of GD in the general population falling between 0.39 to 5.80 per 100,000, and prevalence ranging between 0.70 and 1.75 per 100,000 [19]. The prevalence of GD varies with ethnicity and is markedly higher in Ashkenazi Jewish populations, who also have a higher *GBA* mutation carrier frequency [19]. GD has three clinical subtypes: 1, 2 and 3 [20]. Type 1 GD is the mildest and most common phenotype, accounting for 94% of cases in western populations [19]. Type 1 is characterised by clinicopathological features including bone lesions, hepatosplenomegaly, thrombocytopenia and anaemia, in the absence of primary neurological disease [18]. Types 2 and 3 are neuronopathic forms, with mean lethality at < 4 years old and < 40 years old respectively [21]. *GBA* mutations vary in pathogenicity therefore those that cause type 1 GD (such as N370S) are considered mild, whereas those that also cause type 2 and 3 GD (such as L444P) are denoted as severe [22,23].

### The link Between *GBA* mutations and PD

Within the last three decades, clinicians have reported the presentation of Parkinsonian motor features in a subset of patients with GD [24,25]. An increased incidence of PD within GD kindreds was observed, and PD patients with *GBA* mutations were more likely to report a first or second degree relative with PD, than those without *GBA* mutations [26,27,28]. Parkinsonism has presented in GD patients with both neuronopathic and non-neuronopathic *GBA* variants [29]. Subsequently, in an international multi-centre analysis a strong association was detected between *GBA* mutations and PD, with an odds ratio of 5.43 for carrying heterozygous *GBA* mutations in PD patients vs controls [28]. This work established heterozygous *GBA* mutations as the most common genetic risk factor for PD. Whilst an increased lifetime risk for developing PD has been established for both biallelic and monoallelic carriers of *GBA* mutations, it is still only a minority of individuals with these mutations that will go on to develop PD [30,31]. Heterozygous *GBA* mutation carrier status confers a 3-15% risk of PD by age 80, contingent upon the severity of the variant, however the literature concerning the penetrance of these mutations in PD is variable [23,30,32,33]. Penetrance of *GBA* mutations in PD has been estimated between 10-30% and can be stratified by genetic and nongenetic factors, including age-specificity and severity of mutation [33,34].

#### The prodrome of GBA associated PD

There is an overrepresentation of literature concerning the natural history of PD associated with the two most frequent *GBA* pathogenic variants, L444P and N370S, compared to the role of many of the other hundreds of variants [13,22,35,36,37]. The phenotype of *GBA* associated PD (*GBA* PD), differs from sporadic PD (sPD), with a slightly earlier age of onset of approximately 5 years, with a dose effect for severe vs mild mutations, [38,23]. It has also emerged that *GBA* PD is associated with more non-motor symptoms than its idiopathic counterpart, including a greater likelihood of manifesting cognitive dysfunction concomitant with more aggressive cognitive decline, more severe affective disorder, and an increased risk of experiencing RBD, hyposmia and autonomic dysfunction [39,40,30,41,42].

#### GCase in sporadic PD

Whilst the molecular mechanisms of pathogenesis have not been fully elucidated for *GBA* PD, much like in GD there is a significant reduction in GCase activity [43]. Histopathological analyses of post-mortem PD brain tissue have identified reduced GCase activity and protein levels in regions with elevated expression of α-synuclein [43], and this was subsequently reproduced [44,45]. The most significant reduction in GCase activity was noted in the substantia nigra, and notably these observations were found in both *GBA* PD tissue and tissue from individuals with sPD [43]. Work in iPSC models has suggested that the involvement of GCase in both *GBA* PD and sPD, may result from a bidirectional relationship between reduced lysosomal GCase activity and cytotoxic α-synuclein aggregation [46,47,48]. Due to the implication of deficient GCase activity in both *GBA* PD and sPD, GCase has become an attractive therapeutic target for PD [49,50,51]. Murine and iPSC models of *GBA* PD and sPD have demonstrated the promise of gene therapies and small-molecule chaperone modulation, to increase GCase activity and subsequently reduce α-synuclein accumulation with neuroprotective outcomes [52,53]. Understanding the mechanisms of GCase-mediated PD pathogenesis is therefore a salient research focus for not only *GBA* PD but more widely sPD.

### Objectives and Rationale

The increasing global burden of PD highlights the relevance of utilising PD’s clinical prodrome for earlier diagnosis, and intervention [2]. As the most common genetic risk factor for PD, with premotor features and putative implications for sPD, defining prodromal PD features associated with *GBA* is an important target for enhancing detection of disease and earlier intervention timepoints [28,39,49,3].

The RAPSODI GD study aims to define prodromal symptoms of PD in individuals who carry *GBA* mutations, both homozygote and heterozygote, and to further elucidate the natural history of *GBA* PD. It will test the hypothesis that the presentation of prodromal features of PD for *GBA* can predict the onset of clinical diagnosis. In doing so, it may be possible to develop models for the accurate prediction of individuals at-risk of developing *GBA* PD, affording earlier opportunities for intervention before irreversible neurodegeneration. Secondary objectives of the study include:

1. undertaking PD risk stratification for individuals with biallelic vs monoallelic *GBA* mutations;
2. serving as a platform for future research to identify at-risk individuals for *GBA* PD, to explore biomarkers of *GBA* PD;
3. probing the limited and variable penetrance of the PD phenotype for individuals with *GBA* mutations.

### Design and Methodology

RAPSODI GD is a remote, longitudinal, descriptive, natural history, registry-based study. Repeated measures are delivered on an annual or biennial basis following baseline assessment. Assessment is undertaken via the online study portal (www.rapsodistudy.com), and by post. Saliva sample collection and genotyping for *GBA* and *LRRK2* mutations is an independent measure, occurring once at baseline. The study aims to assess participants for the presence and progression of six prodromal features of PD: 1. Olfactory dysfunction, 2. Motor dysfunction, 3. Cognitive decline, 4. Anxiety and depression, 5. Motor symptoms, 6. REM sleep dysfunction. RAPSODI GD has 25 years of ethical approval from London – Queen Square Research Ethics Committee, (REC reference: 15/LO/1155).

### Sampling

Purposive sampling has been used to recruit participants from the following strata: 1. Heterozygous *GBA* mutation carriers, 2. Homozygous *GBA* mutation carriers, 3. Heterozygote *GBA* mutation carriers with a confirmed clinical diagnosis of PD, 4. Idiopathic PD cases with unknown *GBA* status, 5. Controls. These sampling methods were chosen with the aim of optimising recruitment for each stratum, to develop a large registry of *GBA* mutation homozygotes and heterozygotes. This method seeks to enhance the statistical power necessary for the detection of subtle prodromal features of PD, allowing for future analysis and comparison of the distribution of premotor symptoms of PD between groups.

RAPSODI GD has eight Patient Identification Centres (PICs), where we are partnered with lysosomal storage disorder units, (Royal Free Hospital, Addenbrooke’s Hospital, Queen Elizabeth Hospital Birmingham, Salford Royal Hospital, Cardiff and Vale University Health Board, University College Hospital, Great Ormond Street Hospital, and Manchester Children’s Hospital). From these PICs GD patients with homozygous *GBA* mutations are recruited. These participants act as index cases, to recruit first degree relatives from. These relatives once consented are subsequently screened and determined to be homozygous or heterozygous for *GBA* mutations, or controls. Our sister study, PD FRONTLINE, is a newly established offshoot which screens idiopathic PD cases for *GBA* mutations. All idiopathic PD cases will initially be recruited and screened via the PD FRONTLINE platform to ensure *GBA* mutation status. These *GBA* mutation positive individuals will then be asked to reconsent and reassign from PD FRONTLINE to RAPSODI GD and will act as index cases to contact first degree relatives through. Relatives will be determined to be homozygous or heterozygous for *GBA* mutations, or controls. Family tracing from index cases is only pursued with permission from the participant. Due to the descriptive nature of RAPSODI GD, we envision that recruitment will remain active throughout the full course of the study period, up to the limit of 10,000 participants.

The inclusion criteria for RAPSODI GD are as follows: 1. Homozygous *GBA* mutation carriers, 2. Heterozygous *GBA* mutation carriers, 3. First degree relatives of heterozygous or homozygous *GBA* mutation carriers, 4. Spouses of individuals matching one of inclusion criteria 1-3, 5. Heterozygous or homozygous *GBA* mutation carriers with a confirmed clinical diagnosis of PD, 6. Idiopathic PD cases with unknown *GBA* status, 7. Age 18-90, 8. Informed consent.

The exclusion criteria for RAPSODI GD are as follows: 1. *GBA* mutation negative individuals with a confirmed clinical diagnosis of PD, 2. A history of Parkinsonism, 3. Individuals with other neurological disorders, 4. Individuals on drugs known to be associated with Parkinsonism.

### Remote annual and biennial assessments

Annual remote assessments within RAPSODI GD take place online, via the RAPSODI GD portal (www.rapsodistudy.com). The battery of assessments takes approximately one hour to complete.

#### Informed Consent

Participant consent is gained upon enrolment within the online RAPSODI GD portal (www.rapsodistudy.com), or the PD FRONTLINE portal for participants with idiopathic PD (www.pdfrontline.com).

#### Self-report rating scales

The RAPSODI GD study uses three self-administered scales to assess: 1. Anxiety and depression, 2. Motor symptoms of PD, 3. REM sleep disturbance. Anxiety and depression are examined through the Hospital Anxiety and Depression Scale (HADS) [54]. This 14-item scale determines separate scores for anxiety and depression, between 0-21, distinguishing between mild, moderate, severe and non-cases. Motor symptoms of PD are assessed via the Movement Disorder Society – Unified Parkinson Disease Rating Scale (MDS-UPDRS) part II [55]. This 13-item section of the MDS-UPDRS, rates motor experiences in daily living such as eating and walking, on a scale of 0-4 from normal to severely dysfunctional. REM sleep disturbance is measured via the REM Sleep Behaviour Disorder Screening Questionnaire (RBDSQ) [56]. This 10-item scale assesses clinical features of REM Sleep Behaviour Disorder, such as dream content and sleep movement.

#### Bradykinesia akinesia incoordination test

Motor dysfunction is assessed via the Bradykinesia Akinesia Incoordination (BRAIN) Test [57]. On a standard physical computer/laptop keyboard, participants must alternate between tapping the “s” key and “;” key for 30 seconds per hand. Incoordination, akinesia, dysmetria and kinesia are assigned scores based on the variance in time travelling between the keys, the time spent on keys, the accuracy of pressing the correct keys, and the total number of key presses, respectively.

#### Cognitive testing

Several aspects of cognitive function are assessed via a battery of precision cognitive tests. This was historically delivered via the online platform “CogTrack ™” [58], until the platform expired, and cognitive testing moved to a new platform “NeurOn” in November 2020 (www.neuropsychology.online). The battery includes tasks of image encoding, simple reaction time, digit span backwards, spatial working memory and image recall. Within this battery participants are scored on dysfunction in working and episodic memory, disturbances in attention focus and sustainment, and memory retrieval rate.

#### Hyposmia assessment

Olfactory function is assessed on a biennial basis via the University of Pennsylvania Smell Identification Test (UPSIT) [59]. The UPSIT is delivered remotely through the post, and scores are uploaded directly onto the RAPSODI GD portal by participants. The UPSIT utilises a series of booklets, each containing 10 microencapsulated odorant panels. Participants apply mechanical pressure to the panels to release the odorants, and then select which of four options they believe each odorant to represent. The participants are assigned a score of olfaction sensitivity from 0-40 depending on the accuracy of their responses.

### Genotyping

In their baseline year with RAPSODI GD, participants are posted a saliva sample collection kit (DNA Genotek Oragene.DNA OG-500), with biological substance category B compliant return packaging. Genomic DNA is extracted via the protocol provided in the DNA purification handbook published by DNA Genotek, for use with the Oragene.DNA OG-500 kits [60]. The full 8.9kb amplicon of the *GBA* gene is sequenced for all known deleterious variants, on the Oxford Nanopore MinION, via a protocol validated by Leija-Salazar et al. [61]. Due to RAPSODI GD’s active selection of individuals with *GBA* mutations, there is a likelihood that individuals from an Ashkenazi Jewish population will be overrepresented within the cohort [23]. As such, genomic DNA samples are also MinION sequenced for the most common *LRRK2* mutation, G2019S, as this is a salient and potentially confounding PD associated risk factor for Ashkenazi Jewish individuals [62].

Upon detection of mutation positive cases, DNA samples will be sent for external confirmatory Sanger sequencing in an NHS accredited lab.

### Follow up

The self-reporting scales, Bradykinesia akinesia incoordination test, and cognitive testing will be repeated once a year from baseline. The UPSIT will be repeated once every other year from baseline.

### Future Perspective

The development and validation of clearly defined features of prodromal PD for *GBA* mutation carriers, could eventually allow for the accurate detection of at-risk individuals. Clinical detection of disease in this latent phase, would lend itself to the identification of earlier therapeutic timeframes to deliver neuroprotective interventions before irreversible neurodegeneration can take place. As the mechanisms of pathogenesis in the GCase-mediated pathway of PD are elucidated, biomarkers and therapeutic targets characterized for *GBA* PD may also prove salient in developing preventative measures within sPD as well.

## Data Availability

This is a complete methodology manuscript, therefore no data is reported.

## Notes

**Financial Disclosure:** This study was funded by the MRC (MR/M006646/1), Cure Parkinson Trust, Kattan Trust, an investigator-initiated research grant (IIR-GBR-001110) provided by Shire International GmbH, a member of the Takeda group of companies, and by the joint efforts of The Michael J. Fox Foundation for Parkinson’s Research (MJFF) and the Aligning Science Across Parkinson’s (ASAP). MJFF administers the grant [ASAP-000420] on behalf of ASAP and itself. For the purpose of open access, the author has applied for a CC-BY public copyright licence to the Author Accepted Manuscript (AAM) version arising from this submission. AHVS is supported by the UCLH NIHR BRC.

### Competing Interest Statement

This study was funded by the MRC (MR/M006646/1), Cure Parkinson Trust, Kattan Trust, an investigator-initiated research grant (IIR-GBR-001110) provided by Shire International GmbH, a member of the Takeda group of companies, and by the joint efforts of The Michael J. Fox Foundation for Parkinsons Research (MJFF) and the Aligning Science Across Parkinsons (ASAP). MJFF administers the grant [ASAP-000420] on behalf of ASAP and itself. For the purpose of open access, the author has applied for a CC-BY public copyright licence to the Author Accepted Manuscript (AAM) version arising from this submission. AHVS is supported by the UCLH NIHR BRC.

### Clinical Trial

No trial ID: The manuscript is a research protocol for a prospective study designed to aid in developing a novel diagnostic framework for individuals at risk of developing Parkinson disease, rather than a clinical trial.

### Author Declarations

London, Queen Square Research Ethics Committee (REC reference: 15/LO/1155).

